# Screening and Treatment of Carotid Stenosis in Patients with HPV-Associated Oropharyngeal Cancer

**DOI:** 10.64898/2026.09.01.26361951

**Authors:** Erin K. Greenleaf, Vlad C. Sandulache, Sai P.R. Manikonda, Neal R. Barshes

## Abstract

Patients with history of neck radiation for Human Papilloma Virus (HPV)-associated head and neck cancer experience rapid progression of carotid artery stenosis. The present study sought to determine whether screening and treating asymptomatic carotid artery stenosis in patients with a history of neck radiation is cost-effective. This study is a cost-utility analysis using a probabilistic Markov model over a thirty-year time horizon assessing carotid screening and treatment to avoid neurologic consequences of neck radiation for HPV-associated head and neck cancer. A strategy of no carotid surveillance was associated with a 14.8% cumulative risk of stroke and a strategy of ultrasound surveillance and treatment with TCAR was associated with a 3.0% cumulative risk of stroke. The latter had a median incremental cost of $1.04 million USD and provided a median 39.1 additional QALYs, resulting in a median incremental cost-effectiveness ratio of $26,556 per QALY. In conclusion, this study suggests that ultrasound surveillance and treatment with TCAR for asymptomatic carotid artery stenosis is likely to be cost-effective for patients who have been successfully treated with radiation therapy for HPV-associated head and neck cancer.

---

Surgical intervention for asymptomatic carotid artery stenosis has historically been indicated when stenosis exceeds 70%, provided that the asymptomatic patient has no less than three to five years of expected survival^1^. This pre-operative stipulation for survival is an important tenet of patient selection, as original trial data showed that the benefit realized from carotid intervention in an asymptomatic population is not manifest until several years after the carotid intervention. Concerns for diminished survival among certain patient populations may lead vascular surgeons and other physicians to suspect screening for and treating asymptomatic carotid artery stenosis to be of limited value. Moreover, recent ECST-2 trial data shows that among patients with greater than 50% stenosis and less than 20% five-year risk of ischemic stroke, there is likely no significant benefit of revascularization over optimal medical therapy^2^. Hence, there is mounting evidence to pursue a strategy of non-operative management in patients with asymptomatic and low-risk symptomatic atherosclerotic carotid artery stenosis, particularly in an aged and highly comorbid patient population.

In contrast to atherosclerosis-associated carotid artery stenosis, carotid stenosis that develops secondary to radiation therapy for Human Papillomavirus (HPV)-associated head and neck cancer often occurs in a younger patient population and with different pathophysiologic mechanisms^3–6^. In spite of the relative youth and seemingly unaltered longevity that characterizes these patients, the presence of oropharyngeal squamous cell carcinoma and notions regarding its historically detrimental impact on survival may give vascular surgeons pause when faced with decisions regarding surgical intervention for asymptomatic carotid stenosis.

Furthermore, when weighing risk and benefit of carotid intervention in patients with a history of neck radiation, the higher nerve injury rates following carotid endarterectomy in this patient population may compound one’s reluctance to operate^7,8^. With the advent of transcarotid artery revascularization(TCAR), however, outcomes among patients with a history of neck radiation who are being treated for carotid artery stenosis appear to be promising^9^. Peri-procedural stroke, myocardial ischemia, death and cranial nerve injury have been found to be similar to or lower than that of carotid endarterectomy^10^.

In this manuscript, we report findings from a cost-utility analysis in order to answer the question of whether screening for and treating asymptomatic carotid artery stenosis in patients with a history of radiation for HPV-associated oropharyngeal squamous cell carcinoma is likely to provide clinical benefit and be a cost-effective management strategy.

## Materials and Methods

We used R version 4.1.2 on RStudio desktop to build and run a probabilistic Markov model to simulate clinical events, costs, and health utilities for two management strategies: (1) no screening or treatment of asymptomatic carotid artery stenosis; and (2) ultrasound screening for severe asymptomatic carotid artery stenosis and treatment with TCAR. The primary patient population of interest were 68-year-old men who underwent successful radiation treatment of HPV-associated oropharyngeal cancer one year prior. In the model [Figure 1], all hypothetical patients were assumed to start in the no evidence of disease (NED) clinical state. In any given time cycle, transitions could occur to the following six additional clinical states: (1) cancer recurrence; (2) severe asymptomatic carotid stenosis; (3) underwent TCAR and early (first year post-TCAR) follow-up; (4) late TCAR follow-up (“TCAR-treated”; beyond one year following TCAR); (5) early (first year) after cerebrovascular accident (CVA); (6) later (second year and beyond) after CVA; and (7) death [see Figure 1]. The two TCAR states provided an initial “tunnel” state for the first year status post TCAR because of slightly higher incidence of stroke and mortality, followed by the “TCAR-treated” state in which stroke and mortality generally plateau. Similarly, CVA was modeled in two sub-states: a “tunnel” state for the first year after CVA followed by a second CVA state. This was done to allow the model to distinguish between the higher costs and worse outcomes seen during the first year after CVA versus those seen in years two and beyond^11^. As is conventional and required for Markov models, all patients were modeled as being in only one clinical state at any point in time.

**Figure 1:**
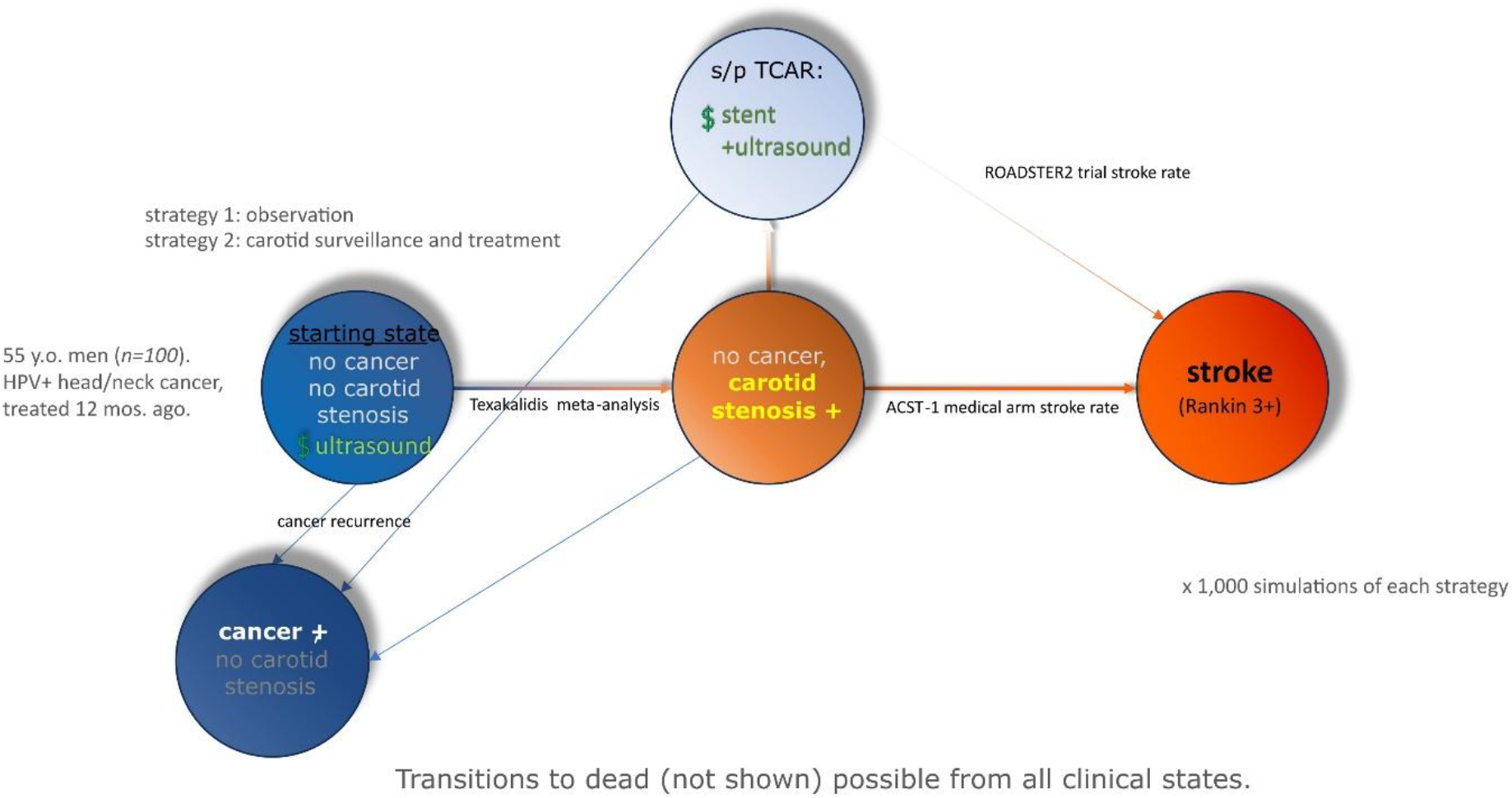
Schematic diagram showing the clinical states simulated in this model.

The probability of various clinical events, the costs, and the health utilities were nearly all obtained from published, peer-reviewed original research. Selected parameters are listed in Table 1 and summarized in the Supplement. We used a 90% estimate of the sensitivity of color duplex ultrasound in detecting a carotid stenosis of 70% or more^12^. The one exception was health utilities with recurrent oropharyngeal cancer, which was modeled as a triangular distribution based on estimates from two (V.S., E.G.) experienced otolaryngologists.

**Table 1.** Selected parameters used in the simulation.

| Variable | Point Estimate | Distribution | Source |
| --- | --- | --- | --- |
| **Stage Transition Probabilities** |  |  |  |
| Probability of successful TCAR for severe carotid stenosis patients in surveillance group | 0.90 | deterministic | ROADSTER * 90% |
| Annual probability of developing carotid stenosis following TCAR | 0.067 | deterministic | Lal et al. |
| TCAR perioperative mortality | 0.002 | deterministic | ROADSTER trial |
| Annual probability of stroke among patients with severe carotid stenosis | 0.023 | deterministic | ACAS trial |
| Annual probability of developing severe carotid stenosis | 0.076 | triangular | Texakalidis et al. |
| Probability of stroke following TCAR | 0.010 | beta | ROADSTER |
| Annual probability of death among patients with no evidence of recurrent cancer | 0.015 | beta | Lop et al. |
| Probability of death within 1 year of stroke | 0.386 – 0.659* | deterministic | Ganesh et al. |
| **Utilities** |  |  |  |
| No evidence of disease | 0.90 | deterministic | best estimate |
| Recurrent cancer | 0.55 | deterministic | best estimate |
| Carotid stenosis | 0.90 | deterministic | best estimate |
| 1<sup>st</sup> year following TCAR | 0.80 | deterministic | best estimate |
| 2+ years following TCAR | 0.85 | deterministic | best estimate |
| 1<sup>st</sup> year following stroke | 0.50 | deterministic | CONTRAST consortium |
| 2+ years following stroke | 0.72 | deterministic | CONTRAST consortium |
| **Costs** |  |  |  |
| TCAR | \$29,455 | gamma | Low, Gray, et al. |
| Screening ultrasound | \$280 | deterministic | Medicare |
| Stroke, 1<sup>st</sup> year | \$76,886 | gamma | CONTRAST consortium |
| Stroke, 2<sup>nd</sup> year | \$19,423 | gamma | CONTRAST consortium |

\* varied with age

The model had a thirty-year time horizon. One thousand simulations were performed for each analysis. All cost values were converted to the March 1st, 2025 United States dollar (USD) values using the “Consumer Price Index Inflation Calculator” from the United States Bureau for Labor Statistics [see https://www.bls.gov/data/inflation_calculator.htm]. We used a 1.5% discount rate for utilities and a 6% discount rate for costs.

Finally, we performed sensitivity analyses for the rate of developing carotid stenosis, as few estimates of this rate have been published for this patient population. These analyses were performed by varying the annual probability of patients in the “no evidence of disease” clinical state developing severe carotid stenosis from 0.5% to 15% per year.

## Results

Simulations projected that the strategy of no surveillance would be associated with a 14.8% cumulative (30 year) risk of stroke, while the strategy of carotid surveillance and treatment would be associated with a 3.0% cumulative risk of stroke [Figure 2 CVAs]. Although the cumulative rate of stroke in the no surveillance group was high, this equated to less than 2% of 100 hypothetical patients being in the “stroke” clinical state (rather than dead, recurrent disease, or with asymptomatic carotid stenosis) at any time in the 30-year time horizon [Supplemental Figure A (state membership)]. Simulations projected that the strategy of carotid surveillance and treatment would require 641 duplex ultrasounds and 94 TCARs per 100 patients over the 30 year model horizon. The number of anticipated TCARs per year decreased from more than 7 per 100 patients in year 2 to approximately 4 per 100 patients in year 10 [Supplemental Figure B (TCARs)].

**Figure 2:**
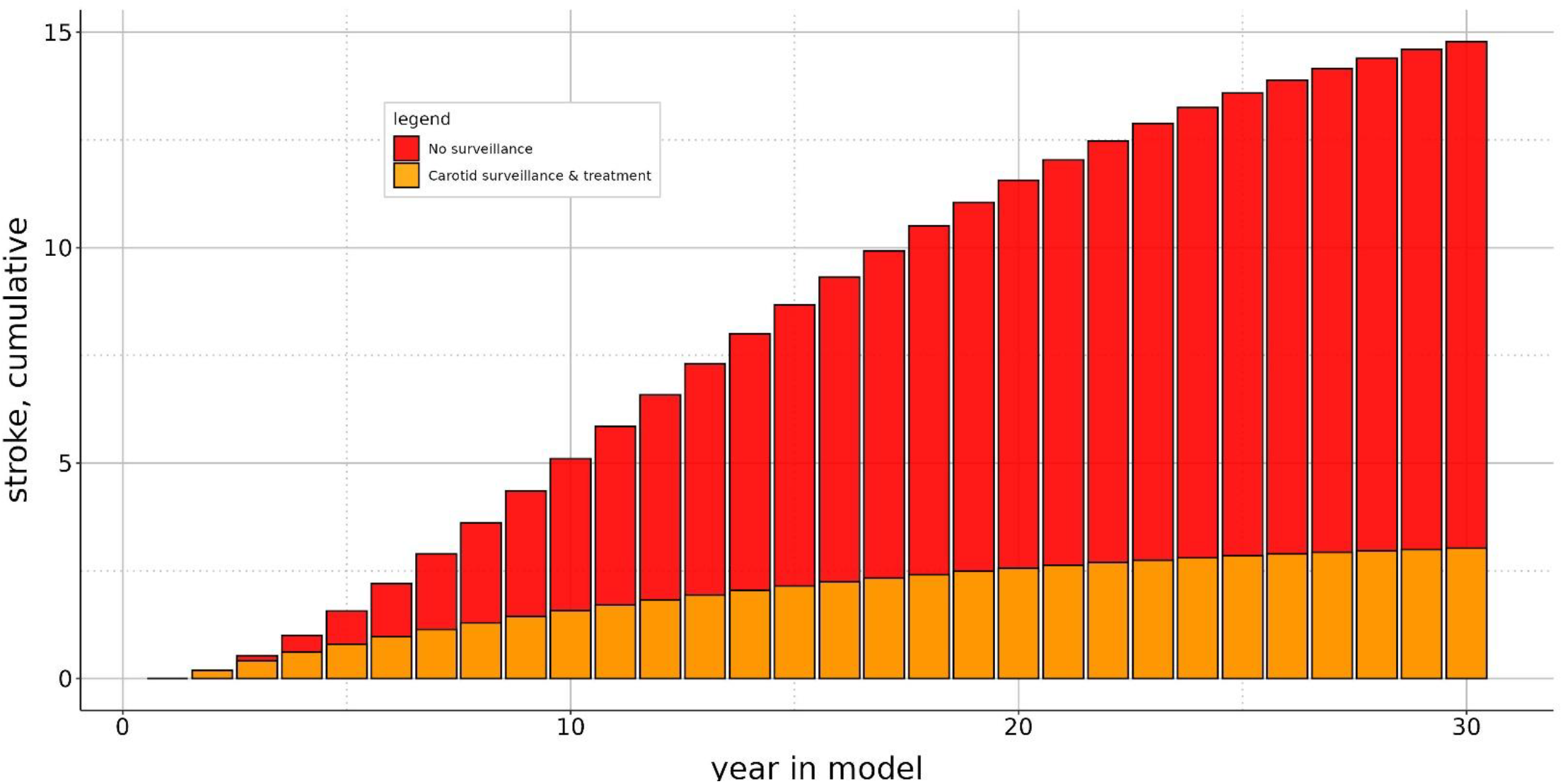
Cumulative risk of stroke according to strategy of surveillance versus strategy of no surveillance.

The strategy of carotid artery surveillance and treatment had a median incremental cost of $1.04 million USD and provided a median of 39.1 additional quality-adjusted life years beyond a strategy of no surveillance. This resulted in a median incremental cost-effectiveness ratio of $26,556 per quality-adjusted life-year. As demonstrated in Supplemental Figure C (Quadrant plot), all simulations projected higher QALYs with the strategy of carotid surveillance and treatment than with the strategy of no surveillance. Many simulations (271 of the 1,000 performed, or 27.1%) projected cost-savings, while 82.0% of simulations were below a willingness-to-pay threshold of $50,000/QALY and 94.4% were below a willingness-to-pay threshold of $100,000/QALY (Supplemental Figure D [CEAC]).

Finally, a sensitivity analysis was performed to understand the impact of variable rates of carotid stenosis -- specifically, annual rates of severe stenosis ranging from 1% to the 7.4% used in the base-case scenario. Sensitivity analyses resulted in ICERs ranging from $18,539 to $24,323 when the annual probability of developing severe carotid stenosis was varied from 0.5 to 15% per year.

## Conclusions

This analysis predicts that a strategy of ultrasound surveillance for and treatment of severe asymptomatic carotid stenosis is likely to be cost-effective -- with possible cost-savings -- for patients who have been successfully treated with radiation therapy for HPV-associated head and neck cancers. A strategy of surveillance remains cost-effective across a wide range of rates of annual progression to severe carotid stenosis. The small, non-linear fluctuations in ICER estimates seen in the sensitivity analysis, wherein the rate of development of severe carotid stenosis was varied, might be attributable to additional costs for TCAR treatment in which the costs of treatment balance the disutility associated with stroke.

The present study adds to the contemporary literature regarding methods of treatment for carotid artery stenosis and their economic value. This cost-effectiveness model assumed carotid stenoses would be amenable to TCAR. The development of TCAR has provided a means for clinicians to treat carotid artery pathology with fewer cranial nerve injuries, lower stroke rate, and lower mortality risk compared to carotid endarterectomy^10,13^. Notably, TCAR has been found to be cost-effective for symptomatic patients within a five-year time horizon relative to carotid endarterectomy^14^.

With regard to screening, most societal guidelines recommend against mass screening programs, but provide qualified recommendations regarding screening in certain high risk populations^4,15,16^. Even in young patients who receive neck radiation for HPV-associated head and neck cancer, longevity is comparable to that of age-matched peers without head and neck cancer^17^. In such populations, preventative medicine remains an important tenet of survival given that these patients largely succumb to either unrelated causes or to the toxicity of cancer treatment. Their risk of developing severe carotid stenosis has been estimated as being higher than that of others because of the neck radiation used in cancer treatment. For these reasons, the strategy of identifying and treating severe asymptomatic carotid stenosis in patients radiated for HPV-associated head and neck cancers would be consistent with societal guidelines, and more specifically, can have a profound impact of the incidence of stroke and the associated disability. The present study adds to the current body of literature insofar as it demonstrates that screening and potentially treating with TCAR is cost-effective relative to an approach of not screening high-risk patients with a history of neck radiation.

The recent publication of the ECST-2 trial data must also be taken into consideration, as one must reconcile the present study’s findings with that of the randomized controlled trial demonstrating no statistically significant benefit of carotid revascularization over optimal medical therapy in patients with asymptomatic and low-risk symptomatic carotid artery stenosis^2^. First, the patient population studied in the ECST-2 trial had carotid artery stenosis as a result of atherosclerosis. The pathophysiology behind this mechanism differs markedly from that of radiation-induced carotid stenosis and demonstrates a different risk profile related to ischemic stroke^18–20^. Second, the Carotid Artery Risk (CAR) score used to stratify patients for the ECST-2 trial has not been validated as a risk prediction tool for those with prior neck radiation and therefore limits the external validity of ECST-2 results for a population with radiation-induced carotid stenosis^21^. Third, the age at which patients with HPV-associated head and neck cancer are diagnosed averages 15 to 20 years younger than the median age of patients in the ECST-2 trial. This gives patients with HPV-associated head and neck cancer significantly more years at risk to not only manifest the sequelae of radiation-induced carotid stenosis but also to aggregate that sequelae with the potential added burden of carotid stenosis related to atherosclerotic risk factors that tend to manifest only later in life. Hence, findings from the ECST-2 trial cannot reasonably be extrapolated to the population of patients treated with neck radiation for HPV-associated head and neck cancers.

There are limitations to this model. First, we incorporated the rates of the patient-level development of carotid stenosis, which seem to consider the development of carotid stenosis in either (left or right) carotid artery. To avoid an overly-complex model, we assumed that only the first carotid artery to develop severe stenosis would be treated and that the other would not progress to severe stenosis. Additionally, the published estimates of the rate of development of severe carotid stenosis following radiation are primarily drawn from patients with head and neck cancers not associated with human papillomavirus -- patients with an older average age and a higher prevalence of tobacco use that would contribute to a background risk of carotid stenosis due to atherosclerotic disease rather than radiation.

Screening for asymptomatic carotid artery stenosis among patients radiated for HPV-associated head and neck cancer appears to be a cost-effective alternative to not screening this population. Future studies should more confidently estimate the incidence of carotid stenosis following treatment of HPV-associated head and neck cancers.

## Data Availability

Data are not publicly available.

## Acknowledgements

We would like to thank Evan Graboyes, MD, MPH, FACS for his intellectual input regarding health utilities in the setting of HPV-associated head and neck cancer.

## Author contributions

EKG and NRB developed the concept for the analysis. NRB curated data, developed the methodology, performed the formal analysis using statistical software. NRB and SPRM created the data visualizations. EKG and NRB wrote the original drafts. NRB, EKG and VCS reviewed and edited the final version.

## Supplemental Materials

**Supplemental Figure A:**
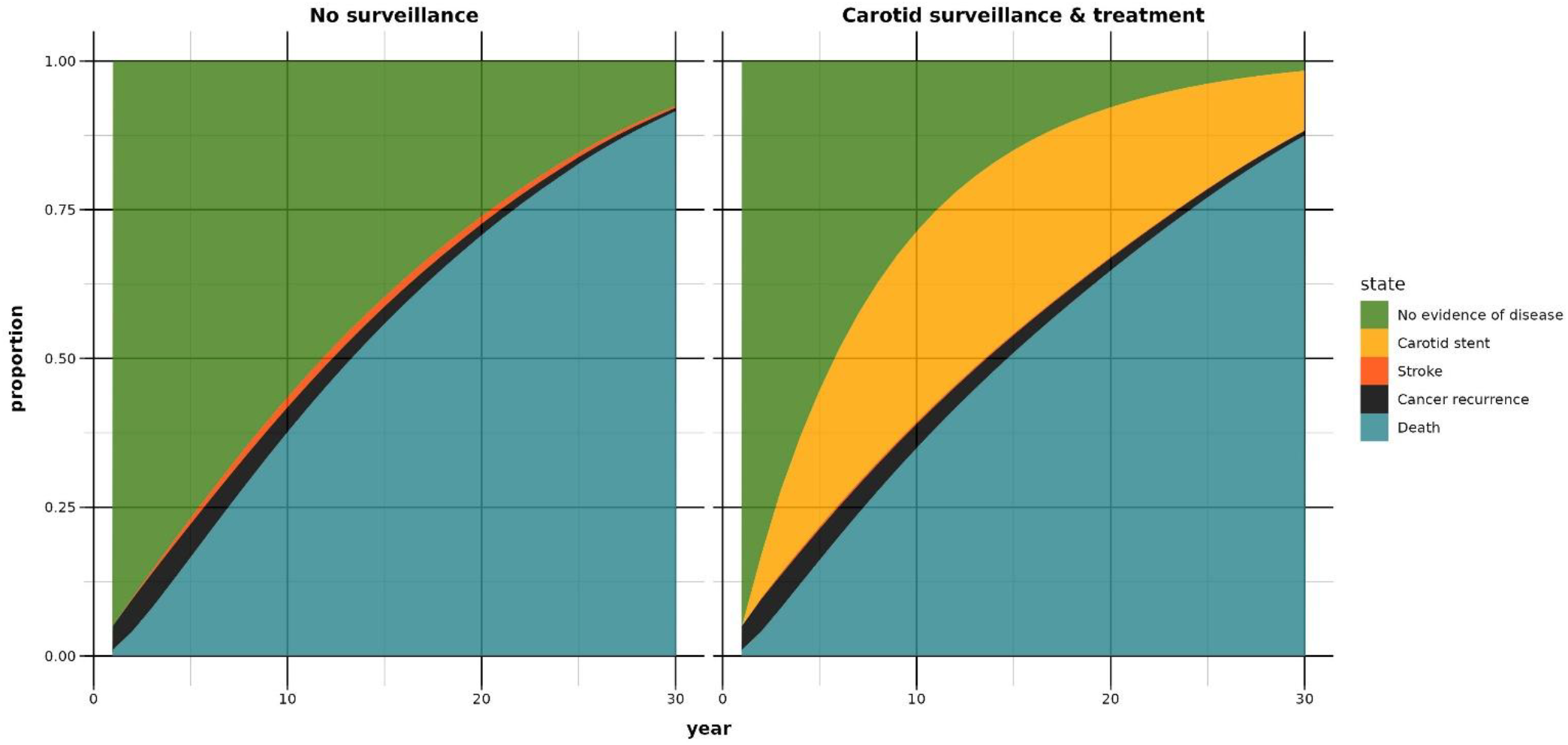
Proportions of patients in each state, stratified by clinical state.

**Supplemental Figure B:**
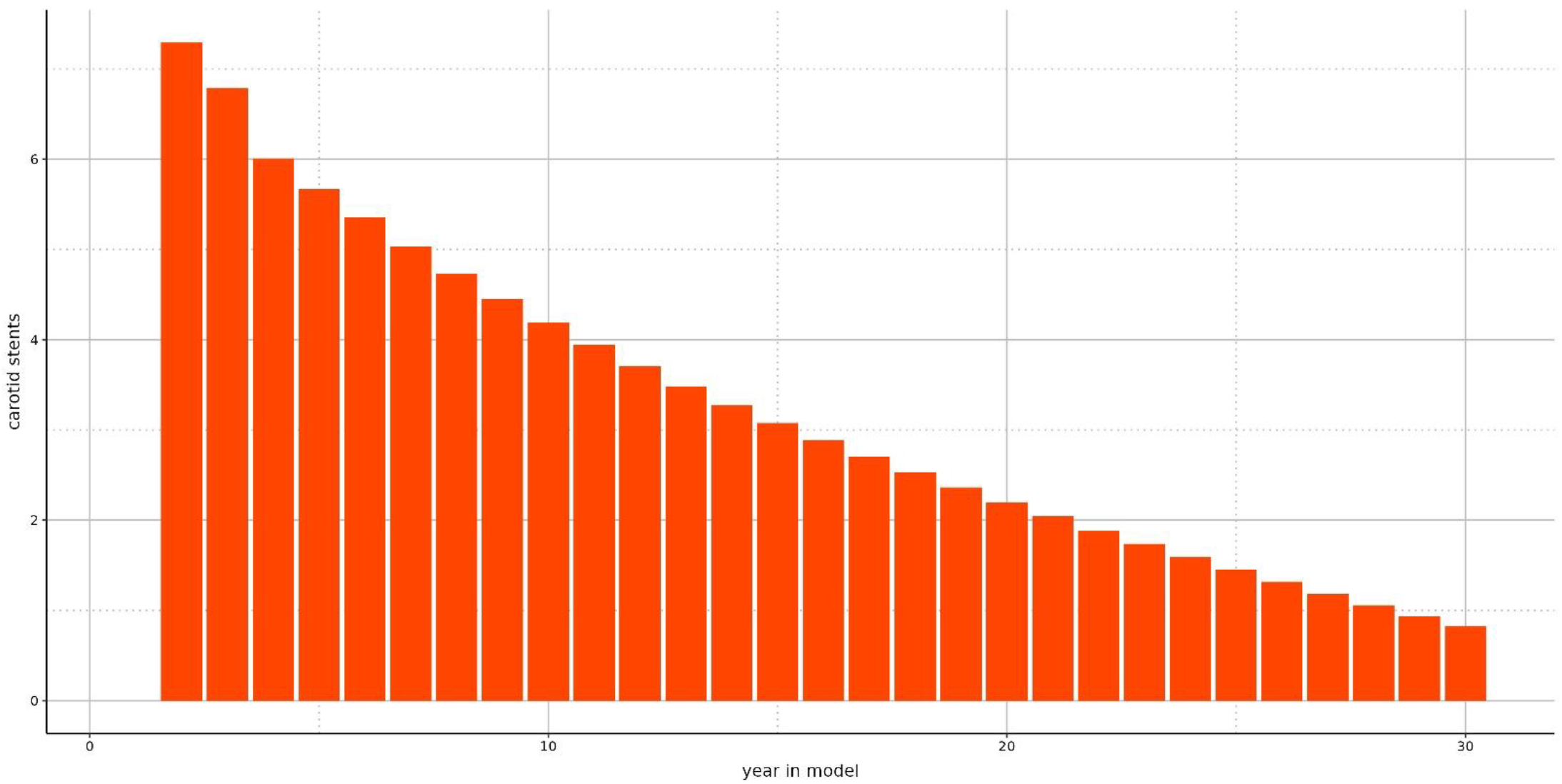
Number of anticipated TCARs indicated over a 30-year time course.

**Supplemental Figure C:**
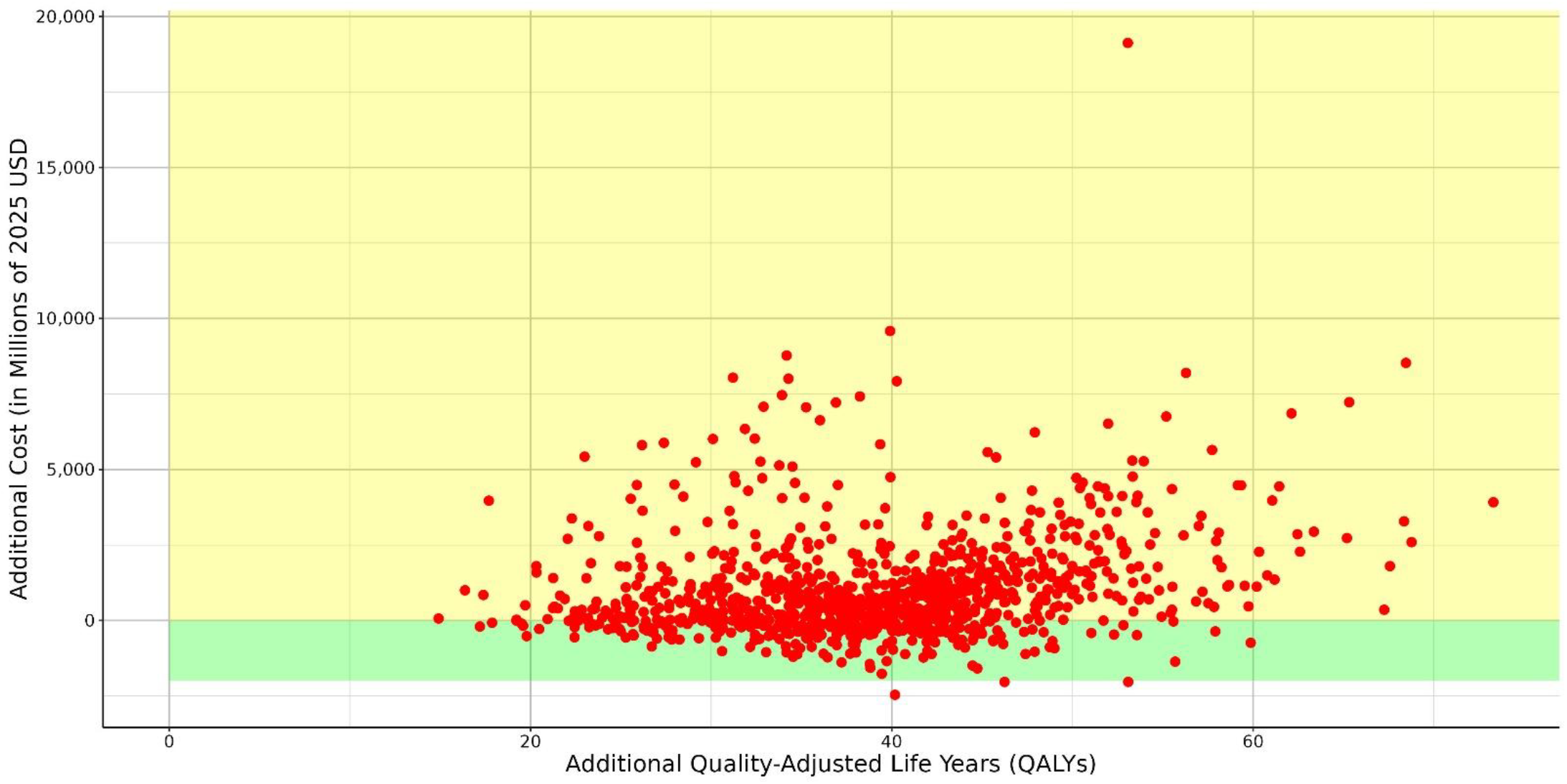
Quadrant plot of QALYs per cost.

**Supplemental Figure D:**
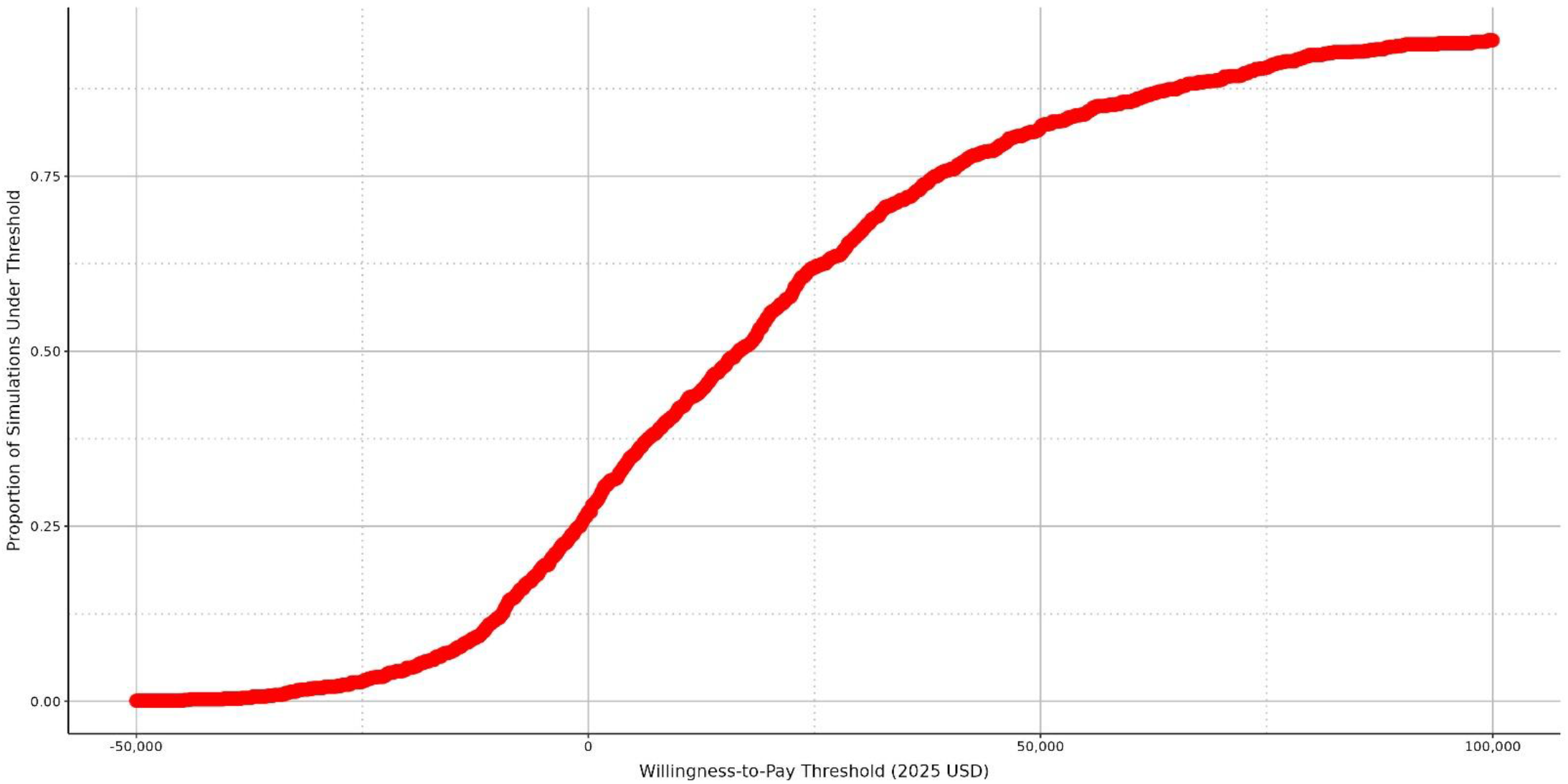
Cost-effectiveness acceptability curve.

